# Examining the Role of Social Environment on COVID-19 Infections and Vaccine Uptake Among Migrants in Japan: Findings from Nationwide Surveys in 2021 and 2023

**DOI:** 10.1101/2024.08.19.24312272

**Authors:** Yuanyuan Teng, Clive E. Sabel, Tomoya Hanibuchi, Tomoki Nakaya

## Abstract

Studies have shown that migrants and ethnic minority groups were unevenly affected by the COVID-19 pandemic, with most evidence coming from Western countries. This study explores the experience of migrants in Japan during the COVID-19 pandemic, examining the association between social environment (i.e., population density, neighborhood deprivation, ethnic density, and social networks) and both COVID-19 infections and vaccination uptake. Data from two nationwide online surveys conducted in 2021 and 2023 were used to analyze these associations during the middle and waning stages of the pandemic. We found that migrants in Japan were more vulnerable to COVID-19 infection but were more receptive to vaccination compared to Japanese nationals. However, not all migrant groups experienced the same outcomes. The effects of social environmental factors on infection and vaccination acceptance were complex and could change, as the pandemic progressed. Environmental factors such as neighborhood population density were not necessarily determinants of COVID-19 infection. More social contacts with Japanese nationals could mitigate vaccination hesitancy, while larger social ties with co-national in the neighborhoods could lead to a higher infection rate. In anticipation of future pandemics, customized programs should be prepared to address the unique healthcare needs of migrants.

## 1. Introduction

The COVID-19 pandemic has had a profound, yet uneven, impact across global populations, with migrants and ethnic minority groups often facing particular vulnerability. Previous studies indicate that ethnic minorities have higher rates of infection, hospitalization, and even COVID-19-related mortality (Duong et al., 2023; Mackey et al., 2021; Magesh et al., 2021; Sze et al., 2020), yet relatively lower vaccine acceptance (Abba-Aji et al., 2022; Khubchandani and Macias, 2021). The disproportionate vulnerabilities among migrants and ethnic minorities stem from socioeconomic disparities, unequal exposure to high-risk occupations, greater pre-existing health challenges, reduced access to healthcare, and lower-quality neighborhood and housing conditions (Duong et al., 2023; Kjøllesdal et al., 2021; Magesh et al., 2021; Parolin and Lee, 2022). However, most of the evidence comes from Western countries, leaving COVID-19 related health outcomes and risk factors among ethnic minorities in Asian contexts, such as Japan, largely under-explored.

Unlike nations with a long history of immigration, Japan has a small but growing foreign population, and its integration policies are limited (Solano and Huddleston, 2020). Despite its non-mandatory infection control measures and its aging society, Japan’s infection and mortality rates of COVID-19 were among the lowest in the world (Amengual and Atsumi, 2021; Iwasaki and Grubaugh, 2020). Given these unique circumstances, the situation faced by migrants and ethnic minorities in Japan may have been more challenging than migrants’ experiences elsewhere.

The first confirmed COVID-19 case in Japan was detected on January 15, 2020. The first Statement of Emergency (SoE) was made on April 7, 2020. Although the SoEs were no mandatory lockdowns, the daily lives of citizens in Japan had largely been restricted. COVID-19 vaccination in Japan began on February 17, 2021, with a staged roll-out to healthcare workers, the elderly, and chronically ill patients, followed by the general population in mid-June 2021 (Kayano et al., 2023). The vaccines were provided free of charge to all residents, including foreigners living in Japan (Fujita et al., 2023). The COVID-19 related health status of ethnic minorities in Japan remains unclear as there are no official data on COVID-19 infection or vaccination rates by nationality or ethnicity. Limited evidence suggests that racial and ethnic minorities in Japan were at a greater risk of COVID-19 exposure although at less risk of severe disease (Nomoto et al., 2022). In addition, no statistically significant changes were confirmed in the mortality differentials between foreign nationals and Japanese citizens for the period from 2015 to 2021 (Ghaznavi et al., 2022).

Due to the less severe Omicron variant and the high rate of vaccination nationwide, the death rate declined since 2021. In early 2022, a new concept entitled "Living with COVID-19" was introduced by the Japanese government, promoting a lifestyle adapted to coexisting with the disease (Kamegai and Hayakawa, 2022). On May 8, 2023, Japan downgraded the legal category of COVID-19 to the same level as common infectious diseases, such as seasonal influenza, marking the beginning of the post-pandemic era. Although the WHO formally declared that the COVID-19 pandemic was over in March 2023, it is still important to understand the health-related situation and its risk factors among migrants in Japan during the COVID-19 pandemic, as new novel threatening viruses could appear anywhere at any time (Biancolella et al., 2023). This study aimed to examine the complex interactions between social factors and both infection and vaccination among foreign residents in Japan, thereby providing valuable insights to better prepare vulnerable groups for possible future global health emergencies.

During the COVID-19 pandemic, social environmental factors have been identified as significant contributors to ethnic health disparities (Finucane et al., 2022; Medcalfe and Slade, 2023; Vahidy et al., 2020; Yang et al., 2021). Living in high population density areas was linked to ethnic differences in COVID-19 infection rates during the early stages of the pandemic (Vahidy et al., 2020). Individuals in deprived neighborhoods faced higher COVID-19 risk and had lower vaccination rates, while ethnic minorities are more likely to live in such neighborhoods due to factors such as financial limitations and discrimination (K.C. et al., 2020; Perry et al., 2021). Studies have also shown that areas with higher ethnic densities experienced greater COVID-19 risk (Yang et al., 2021). Meanwhile, ethnic density could affect vaccination uptake due to factors like concentration of disadvantage of limited vaccination accessibility and political ideologies influencing vaccination intentions (Wu, 2022) Larger social networks might increase COVID-19 risk due to more frequent interactions, but could also enhance vaccination acceptance through prevailing social norms and effective peer pressure. However, this relationship among migrants and ethnic minorities may be unclear as their social networks are relatively limited and greatly impacted by the pandemic. Additionally, as people were less mobile and spent more time in their neighborhoods during the COVID-19 pandemic, social networks within neighborhoods may have had a greater impact on health-related situations than broader social networks.

As social factors can influence health both positively and negatively (Villalonga-Olives and Kawachi, 2017), this study considers the impact of social environment on both infection and vaccination simultaneously. Furthermore, the factors affecting COVID-19 risks may change as pandemics develop due to societal responses (e.g., implementation or lifting disease prevention measures, changes in public compliance), shifting of geographic spread of the virus, the emergence of milder variants, and the introduction of vaccines (Foster et al., 2024; Hoebel et al., 2021; Lundberg et al., 2023; Meurisse et al., 2022; Wu, 2022). Using data from two nationwide online surveys conducted in Japan in 2021 and 2023, this study examined the association of social environment (i.e., population density, neighborhood deprivation, ethnic density, and social networks) with COVID-19 infections and vaccination uptake among migrants in Japan during the middle and the waning stages of the pandemic.

## 2. Materials and methods

### 2.1. Data collection

Since no official data on COVID-19 infection rates and vaccine uptake by nationality or ethnicity is available, we conducted two cross-sectional online surveys among foreign national residents in Japan in 2021 and 2023 to understand their situation during the COVID-19 pandemic. Both surveys were carried out through a survey company named GMO Research, which has access to over 20 million residents in Japan, including more than 20,000 foreign national residents. All foreign registered panelists living in Japan, aged 20 years or older were invited to the survey. Given that online survey panelists tend to have higher levels of education and socioeconomic status (Jang and Vorderstrasse, 2019), and that registering as a panelist requires some knowledge of the Japanese language, it is likely that the participants in our surveys possess higher socioeconomic status and better Japanese language proficiency compared to the average migrant in Japan.

The 2021 survey was conducted from October 5 to October 14, 2021, with questionnaires available in English, Japanese, and plain Japanese (*Yasashii Nihongo*, which uses simpler phrases and fewer Chinese characters). This survey yielded 1,986 valid responses after data cleaning (see Teng et al. (2022) for further details regarding data cleaning). The 2023 survey was conducted from March 24 to 30, 2023, and received a total of 1,063 valid responses. Several modifications were made to the design and implementation of the 2023 survey. First, the questionnaire was only available in English and Japanese. The plain Japanese questionnaire was omitted due to the small number of participants who used it in the 2021 survey and the difficulty of ensuring consistency. Instead, efforts were made to use simpler phrases in the Japanese questionnaire. Second, we limited the number of Taiwanese participants in the 2023 survey because Taiwanese participants in the 2021 survey were over-represented. Third, due to budget constraints, the number of participants in the 2023 survey was capped at 1,100.

Additionally, since the structure and behavior of second-generation migrants in Japan differ significantly from those of the first generation and closely resemble those of Japanese nationals (Teng et al., 2023), this study focuses on first-generation migrants and excluded second-generation migrants from the analysis. Furthermore, participants who did not provide valid postal codes in the questionnaire as well as those where the number of households in the residential area was less than 25 were also excluded to preserve confidentiality. Consequently, the responses used in the analysis are 1,351 and 645 participants from 48 and 33 countries and regions in the 2021 and 2023 survey, respectively.

The study design was approved by the ethics review board of the Center for Northeast Asian Studies of Tohoku University (CNEAS-ER2021-04; CNEAS-ER2022-03). Informed consent was obtained from all participants.

### 2.2. Measures

#### 2.2.1. COVID-19 infection and vaccine uptake

The dependent variable, self-reported COVID-19 infection, was derived from the question on whether the participant ever had been infected with COVID-19 in both surveys. Another dependent variable is vaccine uptake; those who received two or more doses were defined as the fully vaccinated group, while the rest were defined as the incomplete vaccination group. However, given the different progress of vaccination rollout in 2021 and 2023, besides vaccination status, we also included attitude—vaccine hesitancy—as the dependent variable in the 2021 survey. Vaccine hesitancy is defined as a delay in acceptance or refusal of vaccination despite the availability of vaccination services (MacDonald and Hesitancy, 2015). By March 2023, it is reasonable to assume that those who had the intention to be vaccinated would have been. However, in October 2021, many who had not been fully vaccinated were scheduled to be fully vaccinated as the vaccination campaign was still ongoing. Therefore, it is difficult to directly compare vaccination status. In the 2021 survey, the fully vaccinated group and those who were planning to be vaccinated were defined as the intent group. The remaining participants who indicated that they did not want to be vaccinated or had not yet decided were defined as the hesitant group.

#### 2.2.2. Social environment

Social environment was defined to include population density, neighborhood deprivation, ethnic density and social networks. Population density, neighborhood deprivation and ethnic density were calculated using the data from Japan’s 2020 Population Census based on the postal code provided by the participants, while social networks were measured through questionnaires.

Population density was calculated based on the population and area of the postal code area in which the participant resides. Neighborhood deprivation was measured using Area Deprivation Index (ADI), which consists of a weighted sum of eight poverty-related variables including unemployment rate, the proportion of single-mother households, elderly single households, and rented houses. The detailed calculation of the ADI is described in (Nakaya et al., 2014). Higher ADI scores indicate higher levels of deprivation in the neighborhood. Ethnic density was calculated by dividing the foreign populations by the total population of the neighborhood.

#### 2.2.3. Social networks

Regarding social networks, we examined social ties within the neighborhood as well as general social contacts with Japanese.

We distinguished between social ties with co-national neighbors and Japanese neighbors, recognizing that different forms of social capital can have varying impacts on health (Ferlander, 2007). Social capital refers to the resources individuals accessed through their social connections, including networks of relationships, norms of reciprocity and trust. In a migratory context, social ties with co-nationals and locals are usually regarded as bonding and bridging social capital, respectively, based on the similarity of the network (Putnam, 2007, 2000). Bonding social capital refers to connections to people who are like oneself, such as co-nationals, while bridging social capital refers to connections with people who are different, such as native residents to a migrant. To capture these different types of ties, we asked the participants to what extent they agreed with the statement that they know many Japanese/co-nationals in their neighborhood. Options for the 2021 survey included “true,” “somewhat true,” “somewhat untrue,” “untrue,” and “I don’t know”, while options for the 2023 survey used a five-point Likert scale ranging from true to untrue. Questions about social networks were coded as bivariate variables, with those who selected “true” or “somewhat true” as having social ties with Japanese/co-national neighbors, whereas the other answers were coded as other.

Social contacts with Japanese were measured using two items from the social dimension of integration in the Immigration Policy Lab Integration Index (IPL-12) (Harder et al., 2018), which also reflects one’s bridging social capital. The items were “before the outbreak of the COVID-19 pandemic (2021 survey)/ in the last 12 months (2023 survey), how often did you eat dinner with Japanese who are not part of your family?” and “with how many Japanese did you have a conversation - either by phone, messenger chat, or text exchange - in the last 4 weeks?”. Because interpersonal interactions changed tremendously during the pandemic, and the amount of pre-pandemic in person interactions with locals may be more reflective of the reality of social networks, we modified the item in the 2021 survey. Answers to each question were assigned a value from 1 to 5 and summed to form a score. Higher scores indicate more social contacts with Japanese.

#### 2.2.4. Other explanatory variables

In addition to the primary variables of interest, we include several explanatory variables in the models to account for their potential influence. These variables are sex, age, marital status, educational level, work status, household income, length of stay in Japan, home country or region, and residential region. Residential regions of participants were divided into metropolitan and non-metropolitan areas. Metropolitan areas consisted of 11 regions, which are Saitama, Chiba, Tokyo, Kanagawa, Gifu, Aichi, Mie, Kyoto, Osaka, Hyogo, and Nara Prefectures (Osada, 2003). The rest of the regions were coded as non-metropolitan areas. Home country or region were coded into China, Taiwan, Korea, Brazil, Philippines, Vietnam, Western countries, and other.

### 2.3. Statistical analysis

We first calculated descriptive statistics for all variables in the two surveys. Following this, we performed modified Poisson regressions with robust standard errors (Zou, 2004) to examine the association of social environmental factors with COVID-19 infections and vaccination uptake. COVID-19 infection and vaccination experience were the dependent variables, whereas social environmental factors were the independent variables. The ADI was standardized in each survey, and the population density was log-transformed. Crude rate ratios and adjusted rate ratios with 95% confidence intervals were calculated for each coefficient of the model. Statistical analyses were performed using Stata/SE 17.0.

## 3. Results

### 3.1. Descriptive Statistics

The COVID-19 infection experience, vaccination status, and socio-demographic characteristics of the participants in this study are described in Table 1. The participants in the 2023 survey were slightly older than those in the 2021 survey, with mean ages of 40.2 and 37.5 years, respectively. In both surveys, the percentage of female participants was much higher than that of males.

**Table 1.** Descriptive Statistics of participants in two surveys.

|  | 2021<br>(n=1,351) |  | 2023<br>(n=645) |  |
| --- | --- | --- | --- | --- |
|  | n/mean | %/SD | n/mean | %/SD |
| **Self-reported COVID-19 infection** |  |  |  |  |
| No | 1,275 | 94.4 | 403 | 62.5 |
| Yes | 76 | 5.6 | 242 | 37.5 |
| **Vaccination status** |  |  |  |  |
| Fully vaccinated | 450 | 66.7 | 525 | 81.4 |
| Incomplete vaccination | 901 | 33.3 | 120 | 18.6 |
| **Vaccination hesitancy** |  |  |  |  |
| No | 1,196 | 88.5 | - | - |
| Yes | 155 | 11.5 | - | - |
| **Population density (per square kilometer)** | 12054.7 | 8433.5 | 11932.0 | 8284.5 |
| **ADI** | 5.8 | 1.2 | 5.9 | 1.2 |
| **Ethnic density (%)** | 3.9 | 4.4 | 3.9 | 5.9 |
| **Social ties with Japanese neighbors** |  |  |  |  |
| Other | 829 | 61.4 | 370 | 57.4 |
| Yes | 522 | 38.6 | 275 | 42.6 |
| **Social ties with co-national neighbors** |  |  |  |  |
| Other | 1014 | 75.1 | 453 | 70.2 |
| Yes | 337 | 24.9 | 192 | 29.8 |
| **Social contacts with Japanese** | 5.4 | 2.0 | 5.1 | 1.9 |
| **Sex** |  |  |  |  |
| Male | 513 | 38.0 | 242 | 37.5 |
| Female | 838 | 62.0 | 403 | 62.5 |
| **Age** |  |  |  |  |
| 20–29 | 365 | 27.0 | 117 | 18.1 |
| 30–39 | 481 | 35.6 | 204 | 31.6 |
| 40–49 | 305 | 22.6 | 177 | 27.4 |
| 50–59 | 143 | 10.6 | 106 | 16.4 |
| 60 or above | 57 | 4.2 | 41 | 6.4 |
| **Marital status** |  |  |  |  |
| Single | 621 | 46.0 | 258 | 40.0 |
| Married | 730 | 54.0 | 387 | 60.0 |
| **Educational level** |  |  |  |  |
| High school or below | 289 | 21.4 | 122 | 18.9 |
| Higher education | 1,062 | 78.6 | 523 | 81.1 |
| **Work status** |  |  |  |  |
| Regular employee | 597 | 44.2 | 307 | 47.6 |
| Non-regular employee | 378 | 28.0 | 174 | 27.0 |
| Student | 107 | 7.9 | 44 | 6.8 |
| Unemployed/other | 269 | 19.9 | 120 | 18.6 |
| **Household annual income (thousand yen)** |  |  |  |  |
| Below 3,000 | 292 | 21.6 | 129 | 20.0 |
| 3,000–5,000 | 354 | 26.2 | 145 | 22.5 |
| 5,000–8,000 | 260 | 19.3 | 138 | 21.4 |
| Above 8,000 | 187 | 13.8 | 96 | 14.9 |
| Don't know | 258 | 19.1 | 137 | 21.2 |
| **Length of stay in Japan** |  |  |  |  |
| 20 years or more | 429 | 31.8 | 226 | 35.0 |
| 10–19 years | 339 | 25.1 | 176 | 27.3 |
| 3–9 years | 436 | 32.3 | 189 | 29.3 |
| <3 years | 147 | 10.9 | 54 | 8.4 |
| **Home country or region** |  |  |  |  |
| China | 448 | 33.2 | 305 | 47.3 |
| Korea | 215 | 15.9 | 60 | 9.3 |
| Taiwan | 241 | 17.8 | 71 | 11.0 |
| Brazil | 86 | 6.4 | 37 | 5.7 |
| Philippines | 41 | 3.0 | 39 | 6.1 |
| Vietnam | 79 | 5.9 | 29 | 4.5 |
| Western countries | 113 | 8.4 | 46 | 7.1 |
| Other | 128 | 9.5 | 58 | 9.0 |
| **Residential region** |  |  |  |  |
| Non-metropolitan areas | 260 | 19.3 | 157 | 24.3 |
| Metropolitan areas | 1091 | 80.8 | 488 | 75.7 |

SD: standard deviation.
Note: During the statistical analysis, population density was log-transformed, and the ADI was standardized.

In 2022, Japan’s foreign population was nearly 3.1 million, accounting for 2.5% of the total population. Most of the foreign population were from Asian countries (84.4%), with the largest groups coming from China (24.8%), Vietnam (15.9%), South Korea (13.4%), and the Philippines (9.7%). Residents from other regions included Europe (3.0%), Africa (0.7%), North America (2.5%), South America (8.9%), and Oceania (0.5%) (Immigration Services Agency of Japan, 2023). In the 2023 survey, we received a disproportionately large number of responses from Chinese participants (47.3%).

In the 2021 survey, only 5.6% of participants indicated that they had been infected with COVID-19, but in the 2023 survey, this number reached 37.5%. Further, in the 2021 survey, 66.7% of the participants were fully vaccinated, and 21.8% had received a first dose and were planning to receive a second dose. In the 2023 survey, 81.4 % of the participants were fully vaccinated.

In addition, most of our participants resided in metropolitan areas (80.8% in 2021; 75.7% in 2023), consistent with the geographic distribution of the foreign population in Japan (Inoue et al., 2021), although still higher than the official statistics (69.3% in 2022) (Immigration Services Agency of Japan, 2023). Population density, ethnic density, and ADI scores were almost identical between the two surveys.

In terms of social networks, participants reported more social ties in their neighborhoods in 2023 than in 2021, likely indicating increasing in social interactions as the pandemic evolved. Furthermore, because Japanese were the dominant ethnicity in most neighborhoods, participants had more social ties with their Japanese neighbors than with co-national neighbors in both surveys. Furthermore, the 2021 survey results, reflecting pre-pandemic conditions, showed higher scores for social connections with Japanese than the 2023 survey.

### 3.2. Modified Poisson Regression

#### 3.2.1. COVID-19 infection experience

Table 2 presents the results from modified Poisson regression analyses examining the relationship between self-reported COVID-19 infection experience and various social environmental factors. Among these factors, a statistically significant positive association was found only between COVID-19 infection experiences and social ties with co-national neighbors in the 2021 survey. This association persisted even after adjusting for variables such as home country and length of stay in Japan.

**Table 2.**
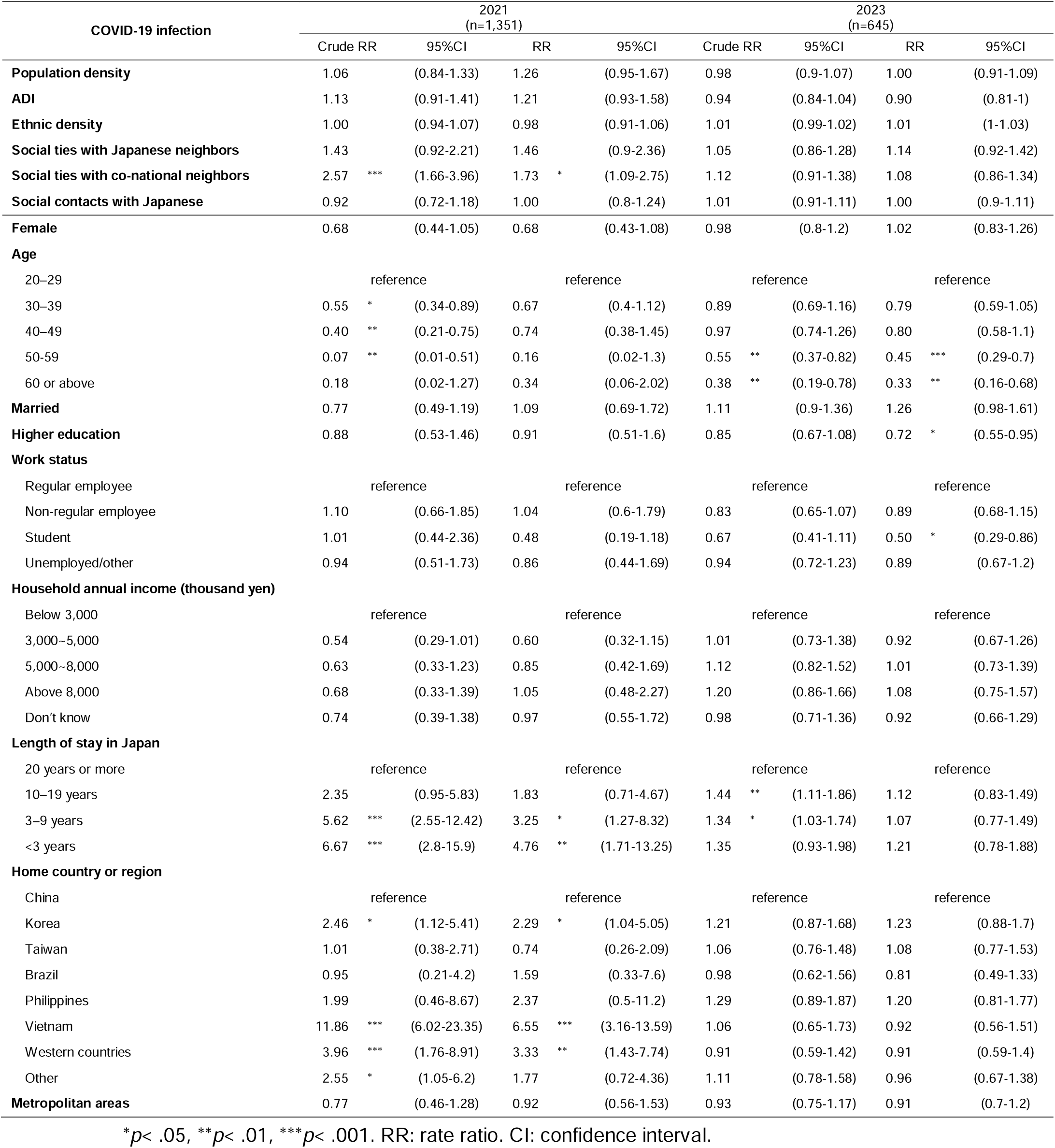
The association between COVID-19 infection and social environment among migrants in Japan.

#### 3.2.2. Vaccine uptake

In the 2021 survey, higher population density was negatively associated with incomplete vaccination before adjustment (Table 3). The ADI was positively related to both incomplete vaccination and vaccine hesitancy before accounting for other variables. Higher ethnic density was negatively associated with incomplete vaccination after adjustments were made. Increased social contacts with Japanese were negatively related to vaccine hesitancy after controlling for other variables. In the 2023 survey, only social contacts with Japanese were significantly related to incomplete vaccination, even after adjustments.

**Table 3.**
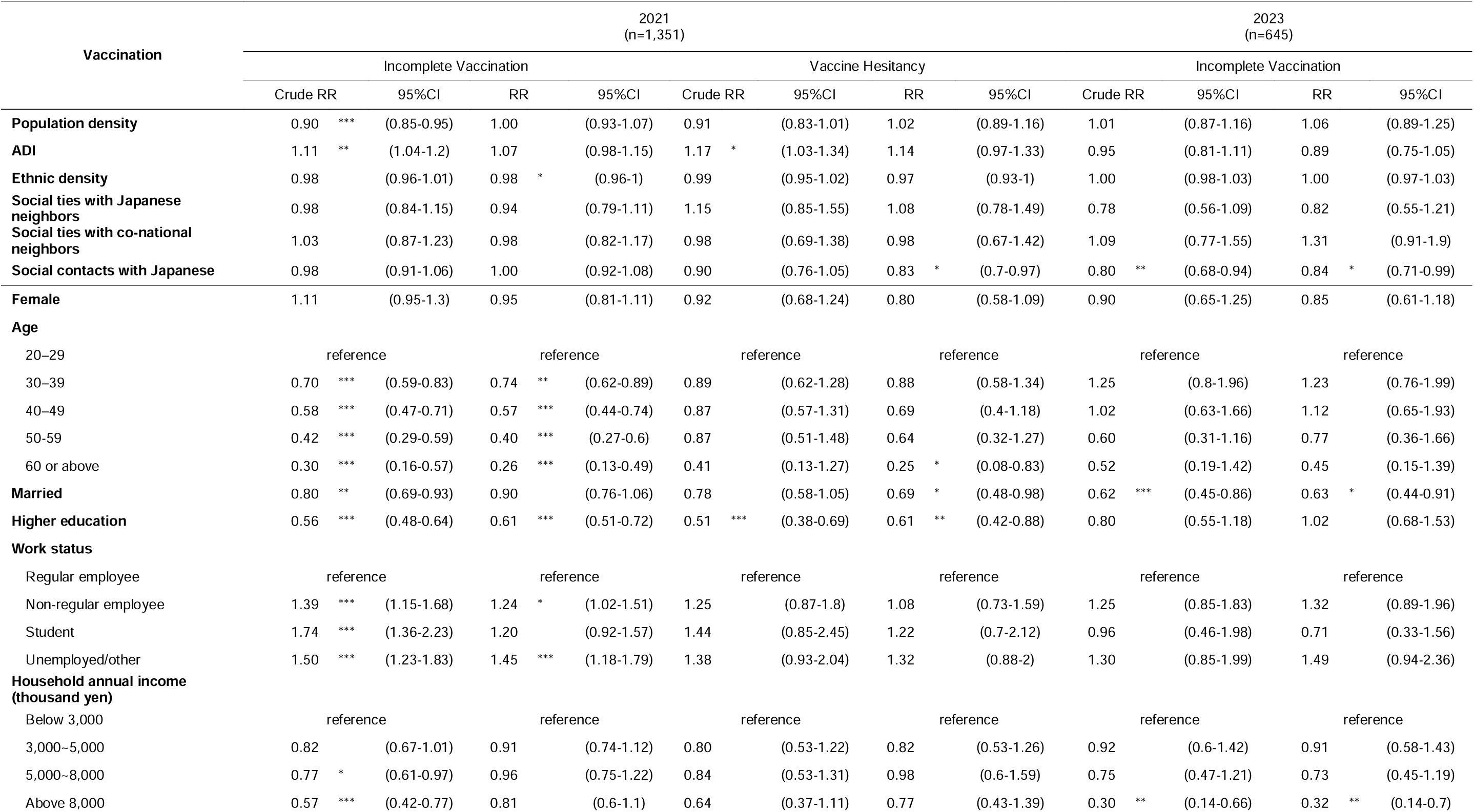
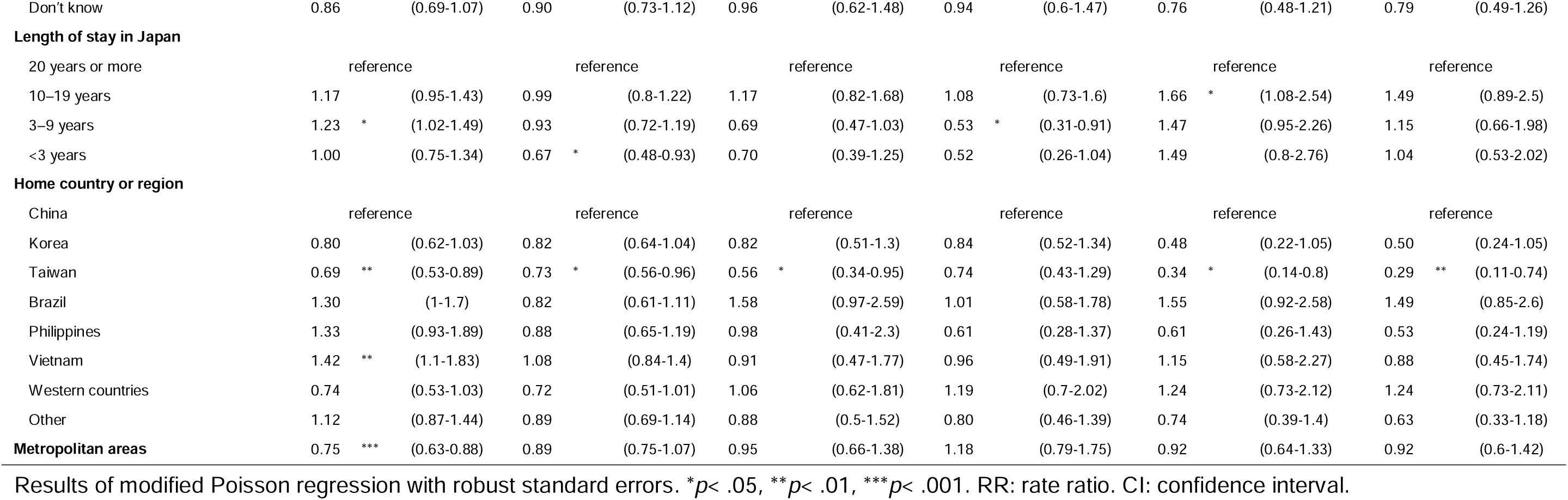
The association between COVID-19 vaccination acceptance and social environment among migrants in Japan.

## 4. Discussion

This study investigated the relationship between social environment and COVID-19 infection and vaccination acceptance among migrants in Japan, comparing the differences between the middle (2021) and waning (2023) stages of the pandemic. We found that the social environmental factors were more associated with COVID-19 infection and vaccination acceptance during the middle stage of the pandemic compared to the waning stage. Additionally, the social environment played a more important role in determining COVID-19 vaccination than infection experiences. Furthermore, our findings suggest that social networks were important determinants of COVID-19 infection and vaccination acceptance, with different forms of social networks exerting varying effects.

Our findings indicate that migrants in Japan were more vulnerable to COVID-19 infection but were more receptive to vaccination compared to Japanese nationals. As of October 4, 2021, and March 23, 2023, there were 1,705,282 and 33,379,221 confirmed COVID-19 cases in Japan, respectively (Ministry of Health, Labour and Welfare, 2023). Excluding repeated infections, the proportions of the population that had a positive COVID-19 test in official data were 1.4% and 26.7%, respectively. While it is not appropriate to directly compare these data with our participants due to age structure differences (migrants tend to be younger), the infection rates self-reported in our survey were substantially higher (5.6% in 2021 and 37.5% in 2023). Although the mortality differentials between foreign nationals and Japanese did not change significantly (Ghaznavi et al., 2022), higher infection rates among migrants could pose a risk to the broader society.

In terms of vaccination, the proportions of the population fully vaccinated in Japan were 69.4% as of October 4, 2021, and 78.1% as of March 23, 2023 (Sapporo Medical University School of Medicine, 2023). These vaccination rates were comparable to those of our survey participants (66.7% in 2021 and 81.4% in 2023). Notably, when considering the age structure differences between migrants and Japanese nationals—with a higher vaccination acceptance rate among the elderly and a younger average age among migrants—migrants showed a higher likelihood of accepting the vaccine compared to Japanese individuals of the same age group. We must acknowledge that different social norms might have existed between host and home countries for migrants. For recent migrants, science knowledge about infection and vaccine efficacy might well have been obtained from the home country not Japan (Gorman et al., 2020). This would have impacted decisions about vaccination and compliance with suggested protective measures.

No statistically significant association was found between COVID-19 infection and either population density or residential region in both surveys, despite higher infection rates in prefectures with greater population density (Kokubun and Yamakawa, 2021). This suggests that the geographic distribution of COVID-19 infection cases among migrants may differ from the overall distribution in Japan. As noted by Alidadi and Sharifi (2023), the relationship between population density and COVID-19 can vary by geographic context. We add that the relationship may also differ among various population groups. However, consistent with the previous studies of general population in Japan (Alidadi et al., 2023) and the U.S. (Kontokosta et al., 2024), population density in smaller geographic scale, such as neighborhood is not necessarily a determinant of COVID-19 infection. One possible explanation is that residents of high-density neighborhoods may feel more threatened by the virus and therefore take more precautions, as suggested by Hamidi et al. (2020) and Kontokosta et al. (2024). Furthermore, unlike ethnic minorities in the U.S. living in densely populated areas with limited ability to modify their mobility behavior (Kontokosta et al., 2024), highly skilled migrants in Japan are more likely to be concentrated in densely populated areas, both regionally and intra-regionally (Kirimura, 2013; Tong, 2019). This allows them more likely to work from home more than migrants in other areas, thus offsetting the risks associated with density. That is, the geographic structure of minority populations varies in different host societies, so the relationship between population density and the risk of infection for minorities will also vary.

Regarding the effect of density on vaccination, incomplete vaccination was significantly and negatively associated with population density and residential region before controlling for other variables in the 2021 survey. However, population density and living in metropolitan areas were not significantly related to vaccine intentions. There are two possible explanations for this. Firstly, as mentioned above, people living in denser areas perceive more risk, which promotes earlier vaccination behavior. Secondly, due to inequalities in regional promotion of vaccinations, densely populated areas and metropolitan areas may have more vaccination sites (e.g., mass vaccination sites set up in big commercial spaces) and better access to transportation.

Regarding neighborhood deprivation, although not significant, the relationship between ADI and COVID-19 infection reversed between 2021 and 2023. Migrants living in deprived neighborhoods were more likely to be infected with COVID-19 in 2021, but less likely to be infected in 2023. This shift may be because, during the middle of the pandemic, migrants in less deprived areas had better housing conditions and a greater ability to work from home, reducing their likelihood of infection. Conversely, as the pandemic waned and milder variants emerged, society returned to normal. Migrants in less deprived areas, more involved in productive activities, may became more susceptible to infection. These results suggest that determinants of infection and vaccination may differ between the mid-pandemic and receding periods.

Furthermore, significant positive relationships were found between neighborhood deprivation and both incomplete vaccination and vaccine hesitancy in the 2021 survey, before accounting for other variables. A plausible explanation for this is that the spread of COVID-19 vaccination-related knowledge in social spaces is uneven, reaching those living in less deprived neighborhoods first. As the vaccination campaigns progress and a majority of the society became vaccinated, increased knowledge dissemination likely promoted vaccination among those in deprived areas.

Concerning ethnic density, no significant association was found between ethnic density and COVID-19 infection, unlike in the case of ethnic minorities in the U.S. (Yang et al., 2021). Previous study indicated that the relationship between ethnic density and the number of infections is stronger in highly segregated areas (Yang et al., 2021). In Japan, due to its small proportion of migrant population, significant residential segregation and uneven ethnic-related geographic distribution of healthcare resources are not as prevalent as in the U.S., which may explain the differing results. Additionally, we found that higher ethnic density was associated with lower incomplete vaccination in 2021. Ethnic communities and religious facilities, such as mosques, played important roles in the early stages of the COVID-19 vaccination rollout in Japan (Kotani et al., 2022; Tamura et al., 2022) providing better access to COVID-19 vaccination for those living in more concentrated areas where ethnic communities or religious facilities tend to form or locate.

In terms of social networks, neighborhood ties were not related to vaccination acceptance, but social ties with co-national neighbors were significantly and positively related to COVID-19 infection in 2021. These results demonstrated that different forms of social capital can have varying impacts on health-related conditions. Migrants who have more social contacts with Japanese, reflecting greater bridging social capital, were more likely to be vaccinated, a finding also noted in our previous study (Teng et al., 2022); this relationship remained evident in 2023. However, geographically proximate bonding social capital may increase infection risk.

Additionally, we found that demographic characteristics and socioeconomic status (e.g., educational levels and work status) were significantly related to health-related situation during the COVID-19 pandemic. Like the general Japanese population (Kayano et al., 2022; Machida et al., 2021), older migrants were less likely to be infected with COVID-19 but more likely to be vaccinated. Married migrants were more likely to receive the vaccine, which may be motivated by protection of family, but they were also more likely to be infected with COVID-19 because their households were crowded than those of single migrants. A higher educational level was significantly related to higher vaccination status and intentions in 2021, and a lower likelihood of COVID-19 infection in 2023. Regarding work status, as Japan adopted a workplace-based COVID-19 vaccination strategy in the early stages of its vaccination campaigns, having a regular workplace or school was significantly associated with earlier vaccination behavior, though not with vaccination intentions. The effect of affiliation on vaccination status diminished as the vaccination campaign progressed.

Lastly, as migrants are a diverse group, we also found that they were affected differently during the pandemic. For example, Vietnamese immigrants, who had higher self-reported infection rate, were vaccinated later than other groups. This disparity might stem from unequal dissemination of COVID-19-related information among ethnic groups. A survey conducted in March 2021, before the vaccine rollout to general population in Japan, among Vietnamese migrants in Japan found that, although over 90% expressed an intention to receive a COVID-19 vaccination, 60% did not know that the COVID-19 vaccination was free (Quy et al., 2021). This highlights a significant information gap among Vietnamese migrants, which likely extends beyond vaccination but to include knowledge about risk factors and protective measures, contributing to a higher likelihood of infection. Regarding the length of stay in Japan, individuals who had lived in Japan for a shorter duration were more likely to be infected with COVID-19, but were less likely to report incomplete vaccination or vaccine hesitancy in 2021. This may be because new migrants maintained stronger connections to their home countries and were more likely to travel between their home country and Japan. This increased their risk of COVID-19 infection while also providing greater motivation to get vaccinated, as vaccination documentation was required for international travel. As the pandemic progressed, increasingly countries loosen the broad control measures and vaccination was not compulsory to enter the country, which may lower the motivation for vaccination for those who travel between countries. Therefore, in 2023, more integrated migrants who had stayed in Japan for a longer period, were more likely to be vaccinated.

Although our study addresses important gaps in the knowledge on health-related situations among migrants during the COVID-19, a few limitations should be mentioned. First, our surveys were conducted through a large online survey panelist network in Japan, which may have led to a selection bias and limited generalizability. Our participants had a certain level of Japanese language proficiency and relatively high socioeconomic status compared to other migrants. In particular, the situation of migrant workers employed in simple labor occupations, particularly in the manufacturing sector, is unlikely to be reflected in our study. Despite this, our participants reported a relatively high infection rate, suggesting that other migrants may be in an even worse situation regarding COVID-19 infection.

It is worth mentioning that while the socioeconomic status of our participants may be relatively high, our results are consistent with a survey of Vietnamese migrants in Japan conducted in January 2022 via Facebook (n=929). In this survey, 32% of the participants were technical interns, a group typically considered to have low socioeconomic status. Despite this, the survey found that 91% of the participants had been vaccinated (National Center for Global Health and Medicine, 2022). However, in another online survey of foreign nationals conducted in May 2023 (n=1042) through users of a job-seeking site, 88.7% of participants had received at least one vaccination, but only 68.9% were fully vaccinated (Kojima, 2024). Moreover, in this survey, only 29.4% had experienced COVID-19 infection, which is lower than our 2023 survey but slightly higher than the infection rate of Japan.

Second, we examined the social environment based on participants’ current residences; however, as migrants are highly mobile population, they are more likely to have changed their residence during the pandemic, which could affect the accuracy of our social environment assessments.

Third, while we aimed to investigate changes in the effect of social factors over time, we did not ask about the specific timing of infection or vaccination. This could make the effects observed in 2023 less clear, as some participants in the 2023 survey may have infected COVID-19 before 2021. However, given that number of COVID-19 cases in 2021 was relatively small compared to 2023 in Japan, it is likely that most participants infected with the COVID-19 between the 2021 survey and 2023 survey.

## 5. Conclusion

Our key finding was that migrants’ COVID-19 experiences in Japan differed from the native Japanese population. While most existing studies on the health-related situations of ethnic minorities during the COVID-19 pandemic come from Western countries, our paper adds understanding of the situation among migrants in an Asian setting. Similar to ethnic minorities in Western countries, migrants in Japan were more vulnerable to infection. However, their vaccination acceptance was not lower than that of the Japanese population. The effects of social environment on infection and vaccination among migrants in Japan may differ from those in Western countries due to differences in the geographic distribution of migrant population and health resources.

The impact of social environment on infection and vaccination was complex and could change as the pandemic progressed. Larger social networks could lead to a higher vaccination risk but also a higher infection rate, according to its form. Importantly, we underscore the importance of expanding general social networks among migrants. While COVID-19 is a transmissible disease, our results indicate that general social networks were not related to infection experiences, but consistently enhanced vaccination uptake throughout the periods studied. The policy implications of this study are clear. Migrants are disproportionality impacted during a pandemic, by a complex set of social and cultural factors, from both the home and destination countries. In anticipation of the next pandemic, plans should be prepared to ensure migrants have targeted schemes to protect their unique healthcare needs.

## Data Availability

All data produced in the present study are available upon reasonable request to the authors.

## Declaration of generative AI and AI-assisted technologies in the writing process

During the preparation of the original draft the first author used ChatGPT in order to improve readability and language. After using this tool, the authors reviewed and edited the content as needed and take full responsibility for the content of the published article.

## Acknowledgements

This work was supported by Promoting Grants for Research Toward Resilient Society 2021 (Tohoku University), Ensemble Grants for Early Career Researchers 2021 (Tohoku University), and the East Eurasian Studies Project of the NIHU Global Area Studies Program.

## References

Abba-Aji, M., Stuckler, D., Galea, S., McKee, M., 2022. Ethnic/racial minorities’ and migrants’ access to COVID-19 vaccines: A systematic review of barriers and facilitators. J Migr Heal 5, 100086. 10.1016/j.jmh.2022.100086

Alidadi, M., Sharifi, A., 2023. Beyond the blame game: Unraveling the complex relationship between density and COVID-19 through a systematic literature review. Cities 141, 104519. 10.1016/j.cities.2023.104519

Alidadi, M., Sharifi, A., Murakami, D., 2023. Tokyo’s COVID-19: An urban perspective on factors influencing infection rates in a global city. Sustain. Cities Soc. 97, 104743. 10.1016/j.scs.2023.104743

Amengual, O., Atsumi, T., 2021. COVID-19 pandemic in Japan. Rheumatol. Int. 41, 1–5. 10.1007/s00296-020-04744-9

Biancolella, M., Colona, V.L., Luzzatto, L., Watt, J.L., Mattiuz, G., Conticello, S.G., Kaminski, N., Mehrian-Shai, R., Ko, A.I., Gonsalves, G.S., Vasiliou, V., Novelli, G., Reichardt, J.K.V., 2023. COVID-19 annual update: a narrative review. Hum. Genom. 17, 68. 10.1186/s40246-023-00515-2

Duong, K.N.C., Le, L.M., Veettil, S.K., Saidoung, P., Wannaadisai, W., Nelson, R.E., Friedrichs, M., Jones, B.E., Pavia, A.T., Jones, M.M., Samore, M.H., Chaiyakunapruk, N., 2023. Disparities in COVID-19 related outcomes in the United States by race and ethnicity pre-vaccination era: an umbrella review of meta-analyses. Front. Public Heal. 11, 1206988. 10.3389/fpubh.2023.1206988

Ferlander, S., 2007. The Importance of Different Forms of Social Capital for Health. Acta Sociol 50, 115–128. 10.1177/0001699307077654

Finucane, M.L., Beckman, R., Ghosh-Dastidar, M., Dubowitz, T., Collins, R.L., Troxel, W., 2022. Do social isolation and neighborhood walkability influence relationships between COVID-19 experiences and wellbeing in predominantly Black urban areas? Landsc. Urban Plan. 217, 104264. 10.1016/j.landurbplan.2021.104264

Foster, T.B., Fernandez, L., Porter, S.R., Pharris-Ciurej, N., 2024. Racial and Ethnic Disparities in Excess All-Cause Mortality in the First Year of the COVID-19 Pandemic. Demography 61, 59–85. 10.1215/00703370-11133943

Fujita, M., Kanda, M., Kiyohara, H., Ikeda, S., Iwamoto, A., Sudo, K., Teshima, Y., Nii, M., Murata, Y., Kato, J., Komatsu, A., Yumino, A., Sawada, T., Sato, H., Nakasa, T., 2023. Migrants’ access to COVID-19 vaccination in Japan: Progress and challenges. J. Migr. Heal. 7, 100169. 10.1016/j.jmh.2023.100169

Ghaznavi, C., Eguchi, A., Tanoue, Y., Yoneoka, D., Kawashima, T., Suzuki, M., Hashizume, M., Nomura, S., 2022. Pre- and post-COVID-19 all-cause mortality of Japanese citizens versus foreign residents living in Japan, 2015–2021. Ssm - Popul Heal 18, 101114. 10.1016/j.ssmph.2022.101114

Gorman, D.R., Bielecki, K., Larson, H.J., Willocks, L.J., Craig, J., Pollock, K.G., 2020. Comparing vaccination hesitancy in Polish migrant parents who accept or refuse nasal flu vaccination for their children. Vaccine 38, 2795–2799. 10.1016/j.vaccine.2020.02.028

Hamidi, S., Sabouri, S., Ewing, R., 2020. Does Density Aggravate the COVID-19 Pandemic? J. Am. Plan. Assoc. 86, 495–509. 10.1080/01944363.2020.1777891

Harder, N., Figueroa, L., Gillum, R.M., Hangartner, D., Laitin, D.D., Hainmueller, J., 2018. Multidimensional measure of immigrant integration. Proc National Acad Sci 115, 201808793. 10.1073/pnas.1808793115

Hoebel, J., Michalski, N., Diercke, M., Hamouda, O., Wahrendorf, M., Dragano, N., Nowossadeck, E., 2021. Emerging socio-economic disparities in COVID-19-related deaths during the second pandemic wave in Germany. Int. J. Infect. Dis. 113, 344–346. 10.1016/j.ijid.2021.10.037

Inoue, T., Koike, S., Yamauchi, M., Ishikawa, Y., 2021. Exploring the impact of depopulation on a country’s population geography: Lessons learned from Japan. Popul Space Place 28. 10.1002/psp.2543

Immigration Services Agency of Japan, 2023. Statistics on foreign residents in Japan 2022. https://www.e-stat.go.jp/stat-search/files?page=1&layout=datalist&toukei=00250012&tstat=000001018034&cycle=1&year=20220&month=24101212&tclass1=000001060399&tclass2val=0 (accessed 6 August 2024) (in Japanese)

Iwasaki, A., Grubaugh, N.D., 2020. Why does Japan have so few cases of COVID-19? EMBO Mol. Med. 12, e12481. 10.15252/emmm.202012481

Jang, M., Vorderstrasse, A., 2019. Socioeconomic Status and Racial or Ethnic Differences in Participation: Web-Based Survey. Jmir Res Protoc 8, e11865. 10.2196/11865

K.C., M., Oral, E., Straif-Bourgeois, S., Rung, A.L., Peters, E.S., 2020. The effect of area deprivation on COVID-19 risk in Louisiana. PLoS ONE 15, e0243028. 10.1371/journal.pone.0243028

Ministry of Health, Labour and Welfare, 2023. COVID-19 trends: Number of confirmed cases (cumulative). https://covid19.mhlw.go.jp/extensions/public/en/index.html (accessed 6 August 2024).

Kamegai, K., Hayakawa, K., 2022. Towards the light at the end of the tunnel: Changes in clinical settings and political measures regarding COVID-19 from 2021, and future perspectives in Japan. Glob. Heal. Med. 4, 327–331. 10.35772/ghm.2022.01071

Kayano, T., Hayashi, K., Kobayashi, T., Nishiura, H., 2022. Age-Dependent Risks of COVID-19 Putatively Caused by Variant Alpha in Japan. Front. Public Heal. 10, 837970. 10.3389/fpubh.2022.837970

Kayano, T., Ko, Y., Otani, K., Kobayashi, T., Suzuki, M., Nishiura, H., 2023. Evaluating the COVID-19 vaccination program in Japan, 2021 using the counterfactual reproduction number. Sci. Rep. 13, 17762. 10.1038/s41598-023-44942-6

Khubchandani, J., Macias, Y., 2021. COVID-19 Vaccination Hesitancy in Hispanics and African-Americans: A Review and Recommendations for Practice. Brain Behav Immun - Heal 15, 100277. 10.1016/j.bbih.2021.100277

Kirimura, T., 2013. Residential Clusters of Foreigners by Nationality in the Twenty-three Wards of Tokyo: Their Socioeconomic Characteristics Related to the Structure of the Residential Areas. Japanese Journal of Human Geography 65, 29–46. 10.4200/jjhg.65.1_29 (in Japanese)

Kjøllesdal, M., Skyrud, K., Gele, A., Arnesen, T., Kløvstad, H., Diaz, E., Indseth, T., 2021. The correlation between socioeconomic factors and COVID-19 among immigrants in Norway: a register-based study. Scand. J. Public Heal. 50, 52–60. 10.1177/14034948211015860

Kojima, H., 2024. Factors Related to COVID-19 Vaccination and Infection Experiences Among Foreign Residents in Japan, in: Secondary Data Analysis Report on the Lives of Foreign Residents Using the “Survey of Foreign Residents” (Time Series Survey). pp. 10-33. https://csrda.iss.u-tokyo.ac.jp/RPS088.pdf (accessed 6 August 2024). (in Japanese).

Kokubun, K., Yamakawa, Y., 2021. Social Capital Mediates the Relationship between Social Distancing and COVID-19 Prevalence in Japan. Inq J Heal Care Organization Provis Financing 58, 004695802110051. 10.1177/00469580211005189

Kontokosta, C.E., Hong, B., Bonczak, B.J., 2024. Socio-spatial inequality and the effects of density on COVID-19 transmission in US cities. Nat. Cities 1, 83–93. 10.1038/s44284-023-00008-2

Kotani, H., Okai, H., Tamura, M., 2022. Mosque as a Vaccination Site for Ethnic Minority in Kanagawa, Japan: Leaving No One Behind Amid the COVID-19 Pandemic. Disaster Med Public 1–3. 10.1017/dmp.2022.78

Lundberg, D.J., Wrigley-Field, E., Cho, A., Raquib, R., Nsoesie, E.O., Paglino, E., Chen, R., Kiang, M.V., Riley, A.R., Chen, Y.H., Charpignon, M.L., Hempstead, K., Preston, S.H., Elo, I.T., Glymour, M.M., Stokes, A.C., 2023. COVID-19 Mortality by Race and Ethnicity in US Metropolitan and Nonmetropolitan Areas, March 2020 to February 2022. JAMA Netw. Open 6, e2311098. 10.1001/jamanetworkopen.2023.11098

MacDonald, N.E., Hesitancy, the S.W.G. on V., 2015. Vaccine hesitancy: Definition, scope and determinants. Vaccine 33, 4161–4164. 10.1016/j.vaccine.2015.04.036

Machida, M., Nakamura, I., Kojima, T., Saito, R., Nakaya, T., Hanibuchi, T., Takamiya, T., Odagiri, Y., Fukushima, N., Kikuchi, H., Amagasa, S., Watanabe, H., Inoue, S., 2021. Acceptance of a COVID-19 Vaccine in Japan during the COVID-19 Pandemic. Nato Adv Sci Inst Se 9, 210. 10.3390/vaccines9030210

Mackey, K., Ayers, C.K., Kondo, K.K., Saha, S., Advani, S.M., Young, S., Spencer, H., Rusek, M., Anderson, J., Veazie, S., Smith, M., Kansagara, D., 2021. Racial and Ethnic Disparities in COVID-19–Related Infections, Hospitalizations, and Deaths. Ann. Intern. Med. 174, 362–373. 10.7326/m20-6306

Magesh, S., John, D., Li, W.T., Li, Y., Mattingly-app, A., Jain, S., Chang, E.Y., Ongkeko, W.M., 2021. Disparities in COVID-19 Outcomes by Race, Ethnicity, and Socioeconomic Status. JAMA Netw. Open 4, e2134147. 10.1001/jamanetworkopen.2021.34147

Medcalfe, S.K., Slade, C.P., 2023. Racial residential segregation and COVID-19 vaccine uptake: an analysis of Georgia USA county-level data. BMC Public Heal. 23, 1392. 10.1186/s12889-023-16235-0

National Center for Global Health and Medicine, 2022. Challenges Faced by Foreigners Regarding Vaccination, Testing, and Living Difficulties (Survey of Approximately 1,000 Vietnamese Residents in Japan). https://kyokuhp.ncgm.go.jp/press_room/press_release/2021/20220309.html (accessed 6 August 2024). (in Japanese).

Sapporo Medical University School of Medicine, 2023. COVID-19 Vaccination Rates by Prefecture. https://web.sapmed.ac.jp/canmol/coronavirus/japan_vaccine.html?a=1 (accessed 6 August 2024). (in Japanese).

Meurisse, M., Lajot, A., Devleesschauwer, B., Cauteren, D.V., Oyen, H.V., Borre, L.V. den, Brondeel, R., 2022. The association between area deprivation and COVID-19 incidence: a municipality-level spatio-temporal study in Belgium, 2020–2021. Arch. Public Heal. 80, 109. 10.1186/s13690-022-00856-9

Nakaya, T., Honjo, K., Hanibuchi, T., Ikeda, A., Iso, H., Inoue, M., Sawada, N., Tsugane, S., Group, J.P.H.C.P.S., 2014. Associations of All-Cause Mortality with Census-Based Neighbourhood Deprivation and Population Density in Japan: A Multilevel Survival Analysis. Plos One 9, e97802. 10.1371/journal.pone.0097802

Nomoto, H., Asai, Y., Hayakawa, K., Matsunaga, N., Kutsuna, S., Kodama, E.N., Ohmagari, N., 2022. Impact of the COVID-19 pandemic on racial and ethnic minorities in Japan. Epidemiology Infect. 150, e202. 10.1017/s0950268822001674

Osada, S., 2003. The Japanese urban system 1970–1990. Prog. Plan. 59, 125–231. 10.1016/s0305-9006(02)00111-3

Parolin, Z., Lee, E.K., 2022. The Role of Poverty and Racial Discrimination in Exacerbating the Health Consequences of COVID-19. Lancet Reg. Heal. - Am. 7, 100178. 10.1016/j.lana.2021.100178

Perry, M., Akbari, A., Cottrell, S., Gravenor, M.B., Roberts, R., Lyons, R.A., Bedston, S., Torabi, F., Griffiths, L., 2021. Inequalities in coverage of COVID-19 vaccination: A population register based cross-sectional study in Wales, UK. Vaccine 39, 6256–6261. 10.1016/j.vaccine.2021.09.019

Putnam, R.D., 2007. E Pluribus Unum: Diversity and Community in the Twenty-first Century The 2006 Johan Skytte Prize Lecture. Scand Polit Stud 30, 137–174. 10.1111/j.1467-9477.2007.00176.x

Putnam, R.D., 2000. Bowling Alone: The Collapse and Revival of American Community. Simon & Schuster, New York.

Quy, P.N., Nam, N.H., An, D.D., Yoshinaka, 2021. The Barrier of COVID-19 Vaccination in Vietnamese living in Japan. https://healthnet.jp/wp-content/uploads/2021/06/75ceb6bd45ba7f7ad3f431e6a086d819.pdf (accessed 6 August 2024). (in Japanese).

Solano, G., Huddleston, T., 2020. Migrant integration policy index 2020. Migration Policy Group.

Sze, S., Pan, D., Nevill, C.R., Gray, L.J., Martin, C.A., Nazareth, J., Minhas, J.S., Divall, P., Khunti, K., Abrams, K.R., Nellums, L.B., Pareek, M., 2020. Ethnicity and clinical outcomes in COVID-19: A systematic review and meta-analysis. Eclinicalmedicine 29, 100630. 10.1016/j.eclinm.2020.100630

Tamura, M., Kotani, H., Katsura, Y., Okai, H., 2022. Mosque as a COVID-19 vaccination site in collaboration with a private clinic: A short report from Osaka, Japan. Prog. Disaster Sci. 16, 100263. 10.1016/j.pdisas.2022.100263

Teng, Y., Hanibuchi, T., Nakaya, T., 2022. Does the Integration of Migrants in the Host Society Raise COVID-19 Vaccine Acceptance? Evidence From a Nationwide Survey in Japan. J Immigr Minor Healt 1–11. 10.1007/s10903-022-01402-z

Teng, Y., Hanibuchi, T., Machida, M., Nakaya, T., 2023. Psychological determinants of COVID-19 vaccine acceptance: A comparison between immigrants and the host population in Japan. Vaccine 41, 1426–1430. 10.1016/j.vaccine.2023.01.037

Tong, Y., 2019. Foreign resident’s characteristics and geographical distribution in Tokyo metropolis: Focusing on highly-skilled professional. Urban Geography 14, 106–114. 10.32245/urbangeography.14.0_106

Vahidy, F.S., Nicolas, J.C., Meeks, J.R., Khan, O., Pan, A., Jones, S.L., Masud, F., Sostman, H.D., Phillips, R., Andrieni, J.D., Kash, B.A., Nasir, K., 2020. Racial and ethnic disparities in SARS-CoV-2 pandemic: analysis of a COVID-19 observational registry for a diverse US metropolitan population. BMJ Open 10, e039849. 10.1136/bmjopen-2020-039849

Villalonga-Olives, E., Kawachi, I., 2017. The dark side of social capital: A systematic review of the negative health effects of social capital. Soc Sci Med 194, 105–127. 10.1016/j.socscimed.2017.10.020

Wu, C., 2022. Racial concentration and dynamics of COVID-19 vaccination in the United States. SSM - Popul. Heal. 19, 101198. 10.1016/j.ssmph.2022.101198

Yang, T.C., Choi, S.E., Sun, F., 2021. COVID-19 cases in US counties: roles of racial/ethnic density and residential segregation. Ethn. Heal. 26, 11–21. 10.1080/13557858.2020.1830036

Zou, G., 2004. A Modified Poisson Regression Approach to Prospective Studies with Binary Data. Am J Epidemiol 159, 702–706. 10.1093/aje/kwh090

